# Maternal health and autism risk: parsing direct and indirect genetic effects using 3-generation family linkage

**DOI:** 10.64898/2026.04.15.26350976

**Authors:** Elias Speleman Arildskov, Vahe Khachadourian, Jakob Grove, Diana Schendel, Stefan Nygaard Hansen, Magdalena Janecka

## Abstract

Despite autism’s prominent genetic etiology and early-life origins, parsing genetic effects contributing to the condition into those that operate directly (via allelic transmission to offspring) vs. indirectly (via influencing prenatal environment) remains challenging. We examined this using a novel design leveraging 3-generation family linkage in Danish national registers.

The cohort included all children born in Denmark from 1998–2015 and their relatives identified through 3-generation family linkage. The analytic sample comprised full maternal cousin pairs, including parallel (children of mother’s sister) and cross cousins (children of mother’s brother). Exposures were diagnoses in the index mother previously associated with offspring autism; the outcome was autism diagnosis in cousins of the index child. We used Cox proportional hazards models to estimate associations separately in parallel and cross cousins, followed by comparisons of these hazard ratios to infer mechanisms.

Several maternal diagnoses (e.g., postpartum hemorrhage, personality disorders, epilepsy) were associated with autism in both parallel and cross cousins, consistent with shared direct genetic effects. Other conditions (e.g., false labor, recurrent major depressive disorder, other anxiety disorders, systemic connective tissue involvement) showed stronger associations in parallel than cross cousins, supporting additional indirect genetic effects operating through the prenatal environment. Adjustment for the same diagnosis in the cousin’s own mother did not substantially change estimates, providing no evidence for an additional role of non-genetic mechanisms associated with the diagnosis.

These findings suggest that both direct and indirect genetic effects contribute to observed links between maternal health and offspring autism, highlighting etiologic heterogeneity and highlighting a registry-based family design to separate these pathways without genetic data.

## Introduction

Autism is a neurodevelopmental condition often diagnosed in childhood based on a dyad of symptoms involving differences in social communications and restrictive/repetitive behaviors^1^. While multiple studies indicate substantial contribution of familial factors to autism etiology – including genetic variation^2,3^ and potential non-genetic factors shared by members of the same family^4^ – the mechanisms through which these factors can impact neurodevelopment remain mostly unknown.

Consistent with the evidence regarding the role of familial factors in autism, we have shown, in a large population-based study, that multiple observational associations between maternal health in pregnancy and autism are largely attributable to shared familial factors — challenging the direct causal effects these maternal conditions may have on the developing fetus^5^. For example, the effects of maternal pre-eclampsia or anxiety disorders were substantially diminished in sibling comparisons, which control for shared familial confounders. Additionally, many of the diagnoses were associated with an increased likelihood of autism in the child regardless of whether the child’s mother or father had the diagnosis during the pregnancy period – even though a paternal diagnosis (*e.g.,* epilepsy, hypertension) has no direct impact on the fetal intrauterine environment — again, indicating that family-level factors correlated with the parental diagnosis, rather than the diagnosis itself, are likely the true cause underlying these associations. These observations echo multiple similar lines of evidence indicating the presence of familial confounding between a wide range of maternal diagnoses - *e.g.,* infertility^6^, infections^7^ or rheumatoid arthritis^8^ – and autism.

While this evidence challenges the causal impact of many maternal health conditions in pregnancy on neurodevelopment, disentangling the exact familial factors underlying these observational associations remains critical for elucidating autism etiology. Such familial factors can span a broad range of influences, including genetic variation and non-genetic factors, with a further distinction across the genetic causes, which can potentially exert their impact on autism through either direct or indirect effects. Here, in line with the literature^9–11^, we define direct genetic effects as those attributable to the genotype of the person in whom the outcome is measured (*e.g.,* the child followed up for an autism diagnosis), and indirect effects as those attributable to someone else’s genotype (*e.g.,* their parent or sibling). Therefore, in the current work, direct genetic effects represent pleiotropic variants associated with both maternal diagnosis and autism; when these variants are transmitted from mother to child, they can impact child’s neurodevelopmental trajectories, reducing or increasing their likelihood of an autism diagnosis. Conversely, indirect genetic effects do not need to be transmitted to the child to exert their effects: in this context, maternal genetic variation impacts the aspects of child’s early environment that are relevant to their neurodevelopment, *e.g.,* maternal folate metabolism. Notably, multiple early-life outcomes and traits like preterm birth or birth weight have already been shown to arise due to a combination of direct and indirect effects^12–14^.

While both direct and indirect genetic effects are shared across family members, the extent of this sharing differs across the relatives. Specifically, indirect genetic effects on the prenatal environment are fully shared between full siblings and maternal half-siblings; 50% shared between cousins whose mothers are sisters (maternal parallel cousins); and not shared between paternal half-siblings or cousins whose mother and father are siblings (cross cousins). While the magnitude of these effects would differ for postnatal influences – which can be exerted by both parents, as well as other persons affecting the child’s environment, irrespective of their relatedness to the child – we assume that autism etiology is primarily traceable to prenatal development, in line with the existing genetic^15–17^, neuroscientific^18,19^, and epidemiologic^20,21^ evidence.

We therefore leverage the extensive, 3-generation pedigree linkage available in the Danish national registers to examine the potential direct and indirect genetic effects contributing to the observational associations between maternal health and autism. We highlight the diverse mechanisms that could underpin the observational effects of different maternal exposures, and offer a new way to examine direct and indirect genetic effects with the use of family data.

## Methods

### Sample

The source population included all children born in Denmark between January 1^st^, 1998 and December 31^st^, 2015, their parents, and grandparents. All individuals were followed up through December 2016. Access to the data was approved by all relevant register authorities, including the Danish Data Protection Agency, Statistics Denmark, and the Danish Health Data Authority.

From the source population, we selected maternal cousin pairs in which both the index child and their cousin were born within the cohort years, regardless of the birth order. We excluded cousin pairs who were related through more than two grandparents.

### Familial relationships

Schematic of the familial relationships used in the analyses is presented in **Figure 1**. “*Index child*” is the child whose mother’s diagnoses are the exposures of interest, and all familial relations (mother, aunt, cousin) are defined in relation to the index child.

**Fig. 1.**
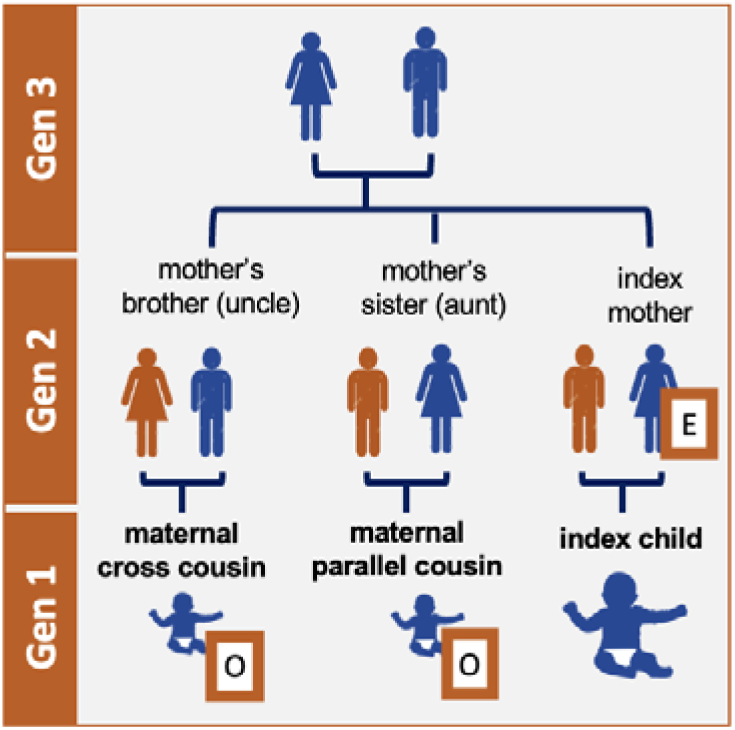
Family relatives in relation to the index child. Dark blue characters indicate blood relatives of the index mother, while orange characters indicate unrelated individuals across 3 generations. “E” indicates the source of the exposure, which is always measured in the index mother. “O” indicates the source of outcome, which is always measured only in maternal cross and parallel cousins of the index child.

All outcomes ascertained in the study are measured in parallel and cross cousins only. In *maternal parallel cousin* pairs, mother and maternal aunt of the index child are full sisters; in *cross cousin* pairs, mother and maternal uncle of the index child are full siblings.

Throughout the analyses, we ascertain only full maternal cousin pairs, *i.e.,* cousin pairs where the mother of the index child and parent of the cousin are full siblings (share both parents).

### Outcome

Outcome of autism was ascertained by a diagnosis of autism spectrum disorder (ASD; ICD-10: F84.0, F84.1, F84.5, F84.8, F84.9 and no diagnosis of F84.2 or F84.3) in the maternal cousins of the index child, obtained via linkage with the Danish Psychiatric Central Research Register (DPCRR)^22^ and Danish National Patient Register (DNPR)^23^. In Denmark, general practitioners or school psychologists refer children with suspected autism to a child and adolescent psychiatric department where they undergo a multidisciplinary evaluation. All ASD diagnoses are given by child and adolescent psychiatrists and are reported according to the International Classification of Disease-version 10 (ICD-10) since 1994; diagnoses from both in- and outpatient contacts have been reported to the DPCRR since 1995. All children in the sample were followed from birth until the first diagnosis of ASD, death, emigration, or end of follow-up (December 31, 2016), whichever occurred first.

### Exposure

We considered diagnoses in the index mother with a previously reported^5^ statistically significant association with the offspring likelihood of autism (**Table S1**). As all outcomes were defined in the cousins of the index child (**Fig. 1**), defining the exposure in their aunt (index mother) enabled us to index a potential genetic liability, rather than the direct impacts of the diagnosis on the cousins themselves.

As this inferred genetic liability is independent of the timing of the diagnosis, the only limits imposed on the exposure window were to increase the likelihood of ascertainment of the diagnosis (which favors longer exposure periods), while minimizing the differences in this likelihood based on the child’s year of birth. Therefore, the exposure period was defined as the period between 48 months preceding birth of the index child until the end of follow-up (Dec 31st, 2016), ensuring a minimal exposure period of 5 years.

To ensure reliable coefficient estimation, we restricted the analyses to the diagnoses with at least 15 exposed and unexposed cross and parallel cousins.

### Covariates

To account for potential confounders, we adjusted for a number of factors related to the index mother (impacting likelihood of exposure) as well as the cousin and aunt (impacting likelihood of outcome).

*Index mother’s covariates* included healthcare utilization, defined as the number of distinct days in the 12 months prior to the birth of the index child during which index mother received any diagnosis (included in the model as a categorical variable with values: 0, 1-3, 4-9 and 10+).

*Cousin/aunt covariates* included (i) cousin’s year of birth, which was included as strata in the model, allowing for individual baseline hazard for each year of birth, in order to adjust for the temporal changes in autism diagnosis; (ii) sex of the cousin, to account for differences in rates of autism diagnosis between males and females; (iii) aunt’s age at cousin’s birth, using restricted cubic splines with 4 knots, to account for increased rates of autism among offspring of very young and older mothers^24,25^; aunt’s (iv) highest attained education and (v) income (using restricted cubic splines with 4 knots) in the year before birth of the cousin, to account for socioeconomic factors influencing the likelihood of the outcome diagnosis in the cousin.

In *<u>additional analyses,</u>* performed only in parallel cousins, we also adjusted for the presence of the exposure diagnosis in the maternal aunt (mother of the parallel cousin). Due to the genetic relatedness between the index mother and her sister, we expect a correlation in their diagnostic status for the exposures used in our study. If these diagnoses affect the child through non-genetic mechanisms, such a correlation could impact the associations in parallel but not cross cousins, invalidating direct comparison of autism risk in these cousin types. Therefore, we verified such potential impact by additionally controlling for the maternal aunt’s diagnosis. The exposure period for the maternal aunt diagnosis was 12 months prior to the birth of the parallel cousin for non-chronic diagnoses, and 48 months prior for chronic ones. Chronicity of the diagnosis was classified in accordance with the Chronic Condition Indicator developed by the Agency for Healthcare Research and Quality^26^. In those models, we also adjusted for the aunt’s healthcare utilization, i.e., the number of distinct days in the 12 months prior to the birth of the index child’s cousin during which the aunt (cousin’s mother) received any diagnosis (as categorical variable, using the division 0, 1-3, 4-9 and 10+).

### Assumptions

All models are based on the following assumptions:

1. There is equal relatedness (shared *<u>direct</u>* genetic effects) between the index child and both their cross and parallel cousins (12.5%).
2. There are shared *<u>indirect</u>*genetic effects, operating prenatally, between the index child and their parallel cousin (whose mothers are full sisters), but not between the index child and their cross cousin (whose mothers are unrelated).
3. *<u>Non-genetic</u>* factors associated with the index mother’s diagnosis include (a) confounders that affect the likelihood of maternal diagnosis and autism (e.g., SES, area pollution), shared to an equal extent with both cross and parallel cousins, and (b) factors operating downstream of the diagnosis (e.g., medication use), which are more likely to be present also in maternal aunt due to diagnostic concordance between sisters. The effects of (a) cancel out when calculating the ratio of hazard ratios between cross and parallel cousins. The effects of (b) are tested in an additional analysis, as explained above.

We also assume that (4) indirect genetic effects operating *postnatally* have no, or little, relevance for autism etiology, consistent with the early prenatal origins of autism^17,18^. While our approach also assumes (5) lack of selection based on differential fecundity in male vs. female siblings of the index mother conditional on her diagnosis (i.e., given the same genetic liabilities); we examine this assumption further by calculating the number of offspring of mother’s brother and sister, stratified by the presence of the diagnosis in the index mother.

### Mechanisms underlying the associations between maternal diagnosis and autism

Using the measures of associations between exposure and risk of outcome in each cousin type, we therefore inferred:

a. Presence of *direct genetic effects* contributing to the association between maternal diagnosis and autism, based on a significant association, in the same direction, between index mother’s diagnosis and risk of outcome the index child, in both cross and parallel cousins.
b. Presence of *indirect genetic effects* operating prenatally contributing to the association between maternal diagnosis and autism, based on a significant association between index mother’s diagnosis and outcome risk in parallel cousins, as well as a significant difference between the coefficients of association of maternal diagnosis with outcome risk between cross and parallel cousins.
c. Presence of *non-genetic* effects contributing to the association between maternal diagnosis and autism, based on a significant reduction in the strength of the association between index mother’s diagnosis and autism in parallel cousins, after accounting for the maternal aunt’s exposure (compared to the model without that adjustment).

These mechanisms (**Fig. 2**) were considered non-mutually exclusive, with each diagnosis-outcome association potentially arising due to a combination of different factors.

**Fig. 2.**
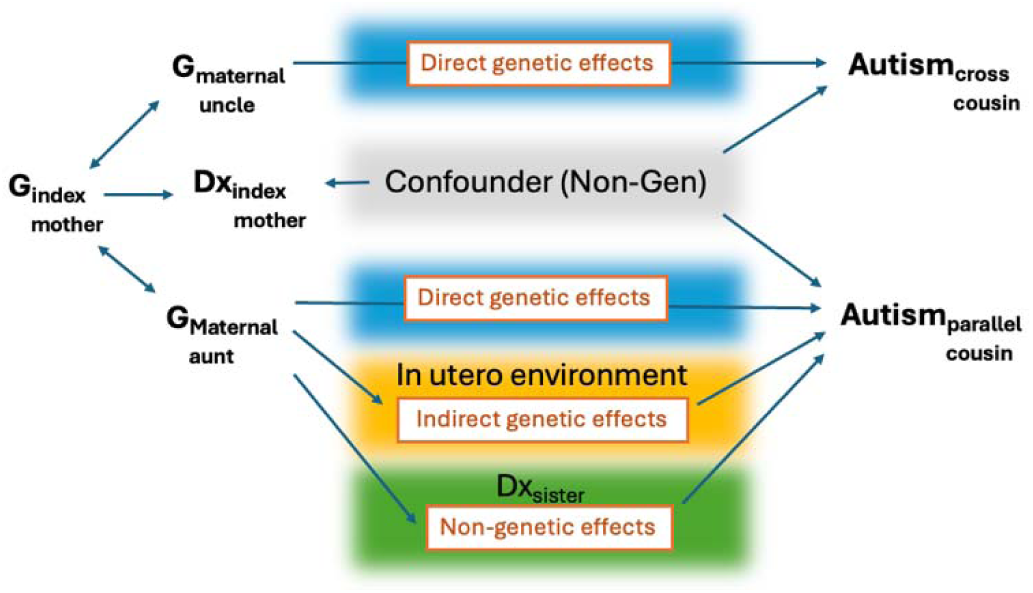
Potential mechanisms contributing to the associations between a diagnosis in the index mother (Dx_index mother_) and autism in cross and parallel cousins of the index child. All familial relationships are in relation to the index child.

### Statistical analysis

We used Cox proportional hazards model to compute the coefficient (log hazard ratio (HR)) of association between each exposure diagnosis (in the index mother) and outcome, separately in each cousin type. Standard errors were calculated for each HR by bootstrapping the entire cousin sample 1000 times. In each bootstrap sample, we computed the coefficients for each cousin type and each diagnosis (i.e., 1000 coefficients per cousin type and diagnosis) and calculated the standard deviation of these 1000 bootstrap coefficients. Based on the coefficients found without bootstrapping (*coef*), and the standard error found by bootstrapping (*SE(coef_b_)*), we calculated p-values (using z-tests) and confidence intervals for the hazard ratios (*95%-CI(HR): exp{coef* ± *z_0.975_SE(coef_b_)}*).

In order to compare the associations of maternal diagnosis with outcome across different cousin types, we computed the ratio of HRs in each cousin type. To obtain the standard errors, we calculated the difference between coefficients for each cousin type in each bootstrapped samples (*diff_b_*), and defined the standard error of the difference (*SE(diff_b_)*) as the standard deviation of the 1000 bootstrap *diff* parameters. We use these parameters to calculate the ratio between hazard ratios (HRR=exp(diff)), p-values (using z-tests) and confidence intervals (*95%-CI(HRR): exp{diff* ± *z_0.975_SE(diff_b_)}*).

The types of cousin comparisons covered maternal full parallel vs. maternal full cross cousins (as described above). Our assumptions regarding each of these comparisons are presented above. We accounted for multiple testing by applying False Discovery Rate (FDR) correction. Nominal and statistical significance were defined as P-value<0.05 and Q-value<0.05, respectively.

### Sensitivity analyses

In the first sensitivity analysis, we verified the impact of the unequal length of the follow-up between children in the cohort - which influences the exposure period and therefore the likelihood of the cousin being ascertained as exposed. To this end, we re-defined the exposure period as the period between 48 months before and 12 months after birth of the index child, ensuring equal exposure period for all cohort children. Secondly, we repeated all analyses in a subset of the data after exclusion of all families where the mother, father, uncle, or aunt of the index child has a diagnosis of autism.

## Results

The source population included 1,131,899 children.

### Evidence for direct genetic effects in the associations between maternal health and autism

There was a nominally significant association between several diagnoses of the index mother and autism in both cross and parallel cousins, with a consistent direction of the effects in both cousin types – indicating the presence of direct genetic effects. These diagnoses included obstetric (abnormalities of forces of labor; postpartum hemorrhage) and injury codes (dislocation/sprain of foot; poisoning by psychotropic drugs), as well as personality disorders and epilepsy (**Fig. 3, left panel; Table S2**). These obstetric codes in the index mother were associated with a *lower* likelihood of autism in index child’s cousins, as opposed to the positive association between maternal diagnosis and child’s autism likelihood we have previously observed^5^. All of these associations were also statistically significant, except poisoning by psychotropic drugs, which was only nominally significant for parallel cousins.

**Fig. 3.**
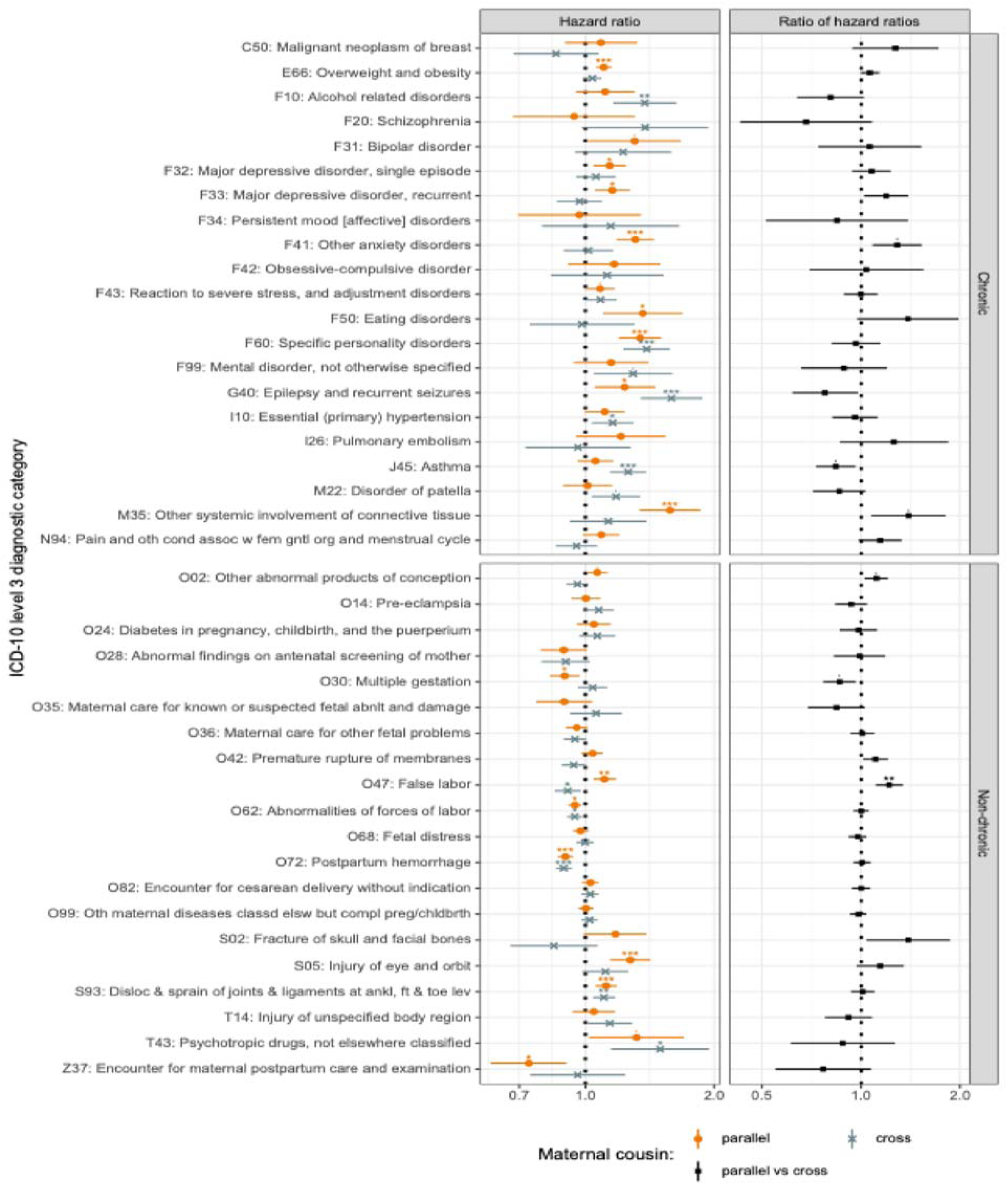
Associations between index mother’s diagnosis and autism in the offspring of her sister (index child’s parallel cousins) and brother (index child’s cross cousins) [left panel] and the ratio of these estimates [right panel].

### Evidence for indirect genetic effects affecting the prenatal environment

Several conditions in the index mother were nominally significantly associated with the likelihood of autism in parallel cousins (children of index mother’s sister), with an effect that was nominally significantly different from that observed in cross cousins (children of index mother’s brother) – indicating presence of indirect genetic effects operating prenatally. These diagnoses included obstetric (abnormal product of conception; multiple gestation; false labor) and psychiatric codes (major depressive disorder (recurrent), other anxiety disorders), as well as systemic involvement of connective tissue (category including e.g., Sjögren and hypermobility syndromes) (**Fig. 3, right panel; Table S2**). All of the conditions were statistically significant associated with autism in the parallel cousins except abnormal product of conception, but the difference between cross and parallel cousins was only statistically significant for false labor.

For all of those diagnoses, the ratio of HR estimates in parallel vs. cross cousins was >1, indicating higher risk in offspring of mother’s sister vs. mother’s brother. The only exception we observed was multiple gestation, which was not associated with autism in children of mother’s brother (cross cousins), but was associated with lower likelihood of autism in children of mother’s sister (parallel cousins) – consistent with the mother-child associations we observed previously^5^.

### Evidence for non-genetic effects due to maternal diagnosis

There was no evidence that the associations between autism likelihood in the cousins and diagnoses in the index mother arose due to non-genetic effects, as defined in our assumptions. Specifically, the estimates of the respective associations remained virtually unchanged after additional controlling for the diagnosis in the cousin’s mother **(Table S3)**.

### Fecundity in family members in relation to maternal diagnosis

Several of the index mother’s diagnoses were associated with changes in fecundity in her siblings. For most diagnoses, fecundity was lower in brothers vs. sisters of the affected females (compared with, respectively, brothers and sisters of unaffected females). These disparities were most pronounced for alcohol use, schizophrenia, bipolar disorder, persistent mood disorders and poisoning by psychotropic drugs (all with >10% difference in relative fecundity). These conditions were associated with normal or higher fecundity in index mother’s sisters, but a substantial decrease in fecundity of her brothers (**Table 1**). Several additional diagnoses were associated with lower fecundity in both brothers and sisters of the affected index mothers, most notably postpartum hemorrhage (over 20% decrease in the number of children), but also other diseases complicating childbirth, OCD, eating and personality disorders. Malignant breast neoplasm and essential hypertension were both associated with higher fecundity in both brothers and sisters of the index mother, likely due to these conditions occurring later in life, when people have attained the highest number of children they were going to have. See **Table S4** for full fecundity information.

**Table 1.**
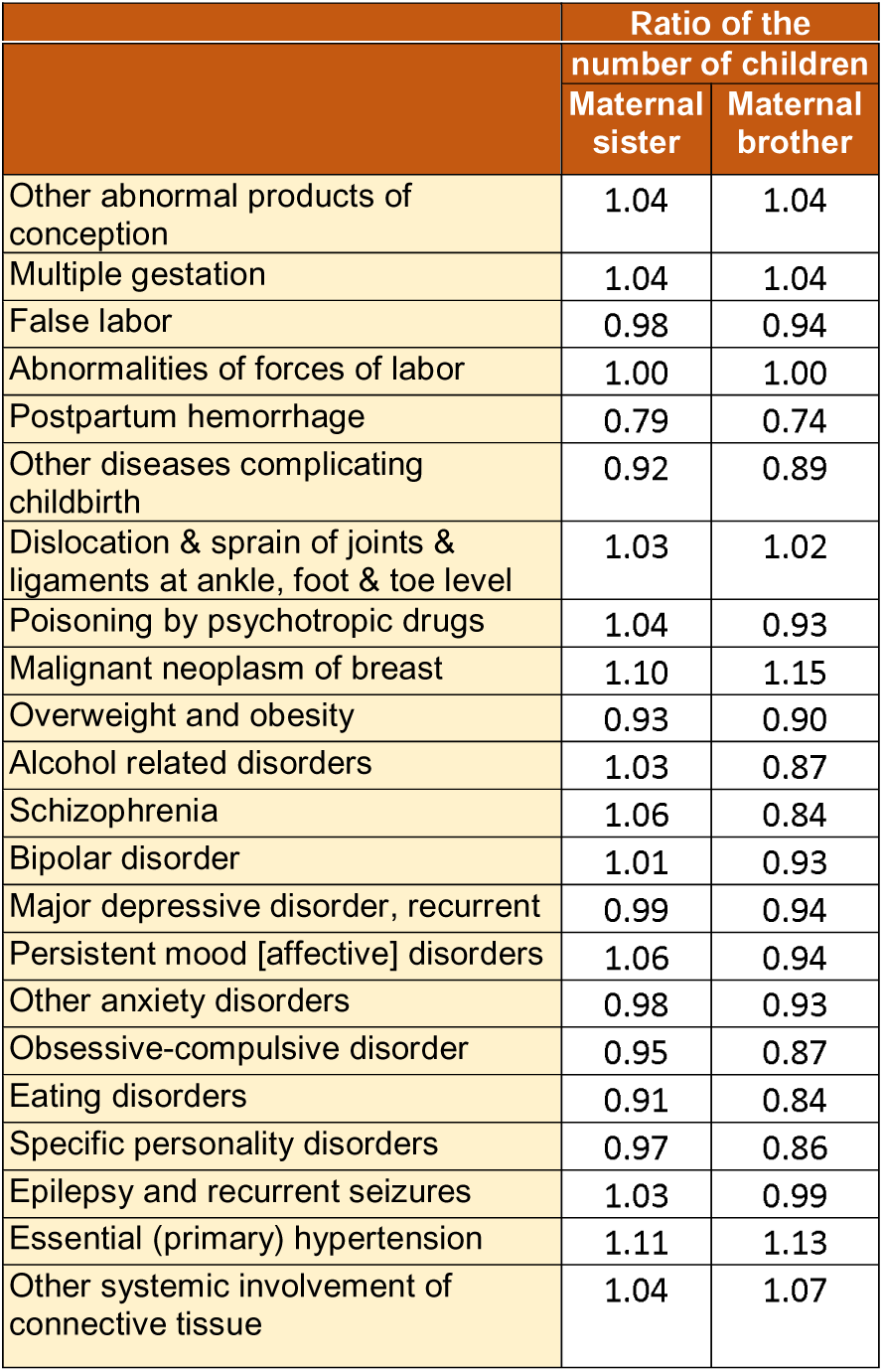
Fecundity in siblings of the index mother, conditional on index mother’s exposure status (selected diagnoses). All ratios were derived by comparing number of offspring in each type of sibling of the index mother with vs. without the given diagnosis (e.g. # offspring sisters of dx(+) index mother / # offspring sisters of dx (-) index mother). Full set of results is presented in Table S4.

### Additional and sensitivity analyses

All associations between index mother’s diagnosis and autism in index child’s cousin remained virtually unchanged after (a) standardizing the exposure period to 5 years across all individuals in the cohort (**Table S5**); and (b) excluding families where any of the index child’s or cousin’s parents had a diagnosis of autism (**Table S6**).

## Discussion

We demonstrated that both direct and indirect genetic effects may contribute to the reported associations between maternal diagnoses around pregnancy and autism in the child, with distinct effects operating across different maternal conditions. These results underscore that the associations between different environmental factors and autism may arise due to heterogeneous mechanisms, fitting in squarely with the notion of complex etiology of autism, and offer new evidence reinforcing the notion of the familial effects contributing to the association between multiple maternal conditions and autism.

Indirect genetic effects represent the effects on one’s outcomes exerted by genotype of another person^10,11^ — typically, through these genotypes affecting outcome-relevant aspects of the index person’s environment (*e.g.,* risk of lung cancer influenced by exposure to second-hand smoke, occurring due to household member’s genetically-influenced propensity to smoke). As such, these effects lie at the intersection of genetic and environmental influences, in opposition to direct genetic effects – which align with the traditional inheritance, whereby one’s outcomes are affected by the genotype they receive from their parents^27^. Delineating between direct and indirect genetic effects is essential for understanding etiology of outcomes with genetic underpinnings, and therefore the pathways to prevention, intervention, and support – as applicable across different traits and conditions.

As both genetic and environmental influences cluster within families, distinguishing between indirect and direct genetic effects is difficult using the established family designs. Presence of indirect genetic effects can be in part inferred from a large attenuation of effect sizes in sibling analyses, as siblings share, on average, 50% of their genetic variation – a ceiling for the attenuation that is attributable to direct genetic effects. Nevertheless, the degree of such attenuation is often hard to estimate given wide confidence intervals around point estimates derived from sibling analyses, as well as the conflation of indirect genetic effects with those exerted by shared environments that are not genetically influenced. Similarly, comparison of the effects of paternal and maternal exposure – potentially useful to delineate the indirect genetic effects operating prenatally, as these are more likely to be maternal in origin – is complicated by the differential exposure rates among males and females, a problem particularly salient when the exposures are health conditions that are strongly sexually dimorphic^28–30^. Therefore, the majority of studies exploring the presence of indirect genetic effects have to date relied on approaches leveraging genetic data, *e.g.,* trio-GCTA^11,31^, trio-PGS^32^ or Genomic SEM^9^.

Indirect genetic effects are of particular interest in the context of autism, given multiple lines of evidence suggesting possible impacts of early-life environment on the likelihood of the condition^18,19^ and the well-established role of genetic variation in neurodevelopment^17^. Here, we were able to elucidate the evidence for indirect genetic effects contributing to the association between child’s autism and maternal obstetric factors (multiple gestation, false labor), affective disorders (major depressive disorder (recurrent), other anxiety disorders) and systemic involvement of connective tissue. These estimates were derived without the genotype data, and avoiding biases due to the differences in the prevalence of those conditions between males and females – as the ascertainment of all diagnoses in our study was done in the index mother only. Our results are consistent with our previous findings indicating that anxiety disorders are associated with lower effects of paternal vs. maternal anxiety on the likelihood of child’s autism (albeit with overlapping confidence intervals between maternal and paternal estimates); and that the effects of recurrent depression, false labor, and systemic involvement of connective tissue are attenuated by >60% in sibling analyses, compared to population-based estimates^5^, *i.e.,* by more than what would be expected based on the sharing of 50% of direct genetic effects between the siblings. Nevertheless, there are also some disparities between the current results and the results from sibling and paternal analyses. For example, for OCD or obesity, we previously observed >80% attenuation of the effects in sibling analyses, yet no significant evidence for the presence of indirect genetic effects in the current approach, and we discuss possible reasons for these discrepancies below. Additionally, other studies exploring the contribution of indirect genetic effects to autism and related traits have found mixed evidence for those^11,33,34^. These disparities may stem from methodological differences; true between-cohort heterogeneity (due to the environmental differences through which indirect genetic effects may operate); or the focus on trait etiology more broadly in the other studies, compared to our consideration of the mechanisms underlying specific observational associations.

Additionally, we observed evidence that the association between a child’s autism and several conditions in their mother – including obstetric (*e.g.,* postpartum hemorrhage) and neuro-psychiatric diagnoses (*e.g.,* personality disorders, epilepsy) – arose due to direct genetic effects. For all of those, we observed that the diagnosis in the index mother was associated with autism in both cross and parallel cousins. These results point towards shared genetic factors between those conditions and autism, with transmission of the relevant alleles to the offspring, and are corroborated by evidence from studies indicating presence of genetic correlations between some of those traits^35,36^. Alternatively, genetic influences associated with the maternal condition could impact fecundity and therefore the levels of selection, with consequences on offspring outcomes – an alternative explanation consistent with e.g., lower fecundity in both male and female siblings of index mothers diagnosed with postpartum hemorrhage, and lower likelihood of autism in their offspring.

While there was no evidence that familial non-genetic effects contributed to the observed associations, this might be due to our inability to distinguish between indirect genetic effects and non-genetic effects, since adjusting for aunt’s diagnosis might not be enough to detect the difference, especially for rarer conditions with only a few exposed aunts.

As highlighted for the association between maternal postpartum hemorrhage and autism, the outcome rates reported in our study could be impacted by certain genetic liabilities on fecundity – with fewer children among the siblings of index mothers with vs. without certain diagnoses. If these effects on fecundity operate in a sex-specific manner, i.e., female and male siblings of the index mother are impacted to a different extent, it could result in differences in autism likelihood between cross and parallel cousins of the index child. Earlier studies have noted that, given the same genetic burden, females tend to have higher fecundity compared to males^37^ – a pattern supported in our sample – translating to higher selection acting on the offspring of males. Therefore, if the genetic factors contributing to those maternal conditions correlate with those for the outcome being measured, this higher selection could lower the outcome rates in the offspring of males. This could account for some of the differences in autism likelihood in cross vs. parallel cousins, conditional on mental health diagnoses in the index mothers, for which we observed highest fecundity differences, and many of which have a strong genetic correlation with autism^2^. Additionally, our assumption of the lack of shared indirect genetic effects acting prenatally between the index child and their cross cousin (whose mother is unrelated to the index child), may be incorrect if there is substantial assortative mating, *i.e.,* if the mother of the cross cousin is “related” to the index family due to non-random mating (demonstrated for a variety of traits and conditions, including psychiatric and cardiometabolic^38^).

The limitations of our study include binarizing the presence of indirect genetic effects based on the nominal significance of the ratio of the estimates in cross and parallel cousins, which may limit our scope to detect those effects if they are more subtle, even in a large, national sample like ours. Additionally, relatedness in our study was based on the registry data and not verified genetically, leading to its potential misclassification, with the risk of misclassification being largest for fathers, who are more likely to not be biological relatives of the index child. Therefore, some of the differences between cross and parallel cousins might not be due to indirect genetic effect only, but to the larger risk of misclassification of cross cousins. Next, if direct and indirect genetic effects act in opposite directions, it may increase the effects in cross vs. parallel cousins, a pattern we observed for e.g., epilepsy and asthma. Further, the families were ascertained into the cohort based on at least one livebirth in the index mother, leading to potential exclusion of families with conditions with the highest impact on fecundity. This is further amplified by the fact that families where either the index mother or her siblings have many children will have a stronger impact on the results than families where the index mother and her siblings have fewer children, since we pair cousins together.

In conclusion, our large, population-based study implemented a novel design that leverages familial relationships across 3 generations of descendants, as well as registry-based clinical and sociodemographic data. Our approach offers a new direction for distinguishing between direct and indirect genetic effects, in the absence of genotype data, and in a manner that is robust to the potential sexual dimorphism of the exposure. Future studies should re-examine the contribution of direct and indirect genetic effects using alternative approaches, and verify the impact of assortative mating on the study results.

## Data Availability

The Danish Scientific Ethical Committee system, as well as all relevant register authorities, including the Danish Data Protection Agency, Statistics Denmark, and the Danish Health Data Authority, approved access to these data under strict conditions regarding access and data export. Under these conditions there are no provisions for exporting individual level data, all or in part, to another institution, in or outside of Denmark. All summary statistics of the measures of associations used to draw the study conclusions are presented in the supplemental material. The corresponding author will respond to any potential additional queries.

## Acknowledgements

This work was supported by grants from the National Institute of Mental Health (MH124817, T32-MH122394) and the Lundbeck Foundation (iPSYCH, grant nos. R102-A9118 and R155-2014-1724).

## Conflict of interests

VK is currently employed by Takeda Pharmaceuticals, outside of the submitted work.

## Supplemental material

**Supplemental Table 1.** Index mother’s diagnoses ascertained in the study. . The chronic/non-chronic status was relevant only in additional analyses in parallel cousins controlling for maternal aunt’s (i.e. cousin’s mother’s) diagnostic status.

| ICD-10 code | Description diagnosis | Chronic / non-chronic status in additional analyses (aunt's status) |
| --- | --- | --- |
| O02 | Other abnormal products of conception | Non-chronic |
| O14 | Gestational hypertension with significant proteinuria | Non-chronic |
| O24 | Diabetes mellitus in pregnancy | Non-chronic |
| O28 | Abnormal findings on antenatal screening of mother | Non-chronic |
| O30 | Multiple gestation | Non-chronic |
| O36 | Maternal care for other known or suspected foetal problems | Non-chronic |
| O42 | Premature rupture of membranes | Non-chronic |
| O47 | False labour | Non-chronic |
| O68 | Labour and delivery complicated by foetal stress [distress] | Non-chronic |
| O99 | Other maternal diseases classifiable elsewhere but complicating pregnancy, childbirth and the puerperium | Non-chronic |
| S02 | Fracture of skull and facial bones | Non-chronic |
| S05 | Injury of eye and orbit | Non-chronic |
| T14 | Injury of unspecified body region | Non-chronic |
| T43 | Poisoning by psychotropic drugs, not elsewhere classified | Non-chronic |
| C50 | Malignant neoplasm of breast | Chronic |
| E66 | Obesity | Chronic |
| F10 | Mental and behavioural disorders due to use of alcohol | Chronic |
| F20 | Schizophrenia | Chronic |
| F21 | Schizotypal disorder | Chronic |
| F31 | Bipolar affective disorder | Chronic |
| F32 | Depressive episode | Chronic |
| F33 | Recurrent depressive disorder | Chronic |
| F41 | Other anxiety disorders | Chronic |
| F42 | Obsessive-compulsive disorder | Chronic |
| F43 | Reaction to severe stress, and adjustment disorders | Chronic |
| F50 | Eating disorders | Chronic |
| F60 | Specific personality disorders | Chronic |
| F99 | Mental disorder, not otherwise specified | Chronic |
| G40 | Epilepsy | Chronic |
| I26 | Pulmonary embolism | Chronic |
| J45 | Asthma | Chronic |
| M22 | Disorders of patella | Chronic |
| M35 | Other systemic involvement of connective tissue | Chronic |
| N94 | Pain and other conditions associated with female genital organs and menstrual cycle | Chronic |

**Supplemental Table 2.** Associations between index mother’s diagnosis and autism risk in maternal parallel and cross cousins (HR, 95% CI), as well as the ratio of these estimates. Associations that were nominally significant in the main analyses are indicated in bold.

|  |  | Maternal parallel cousins |  |  | Maternal cross cousins |  |  | Estimate's ratio<br>(parallel / cross) |  |  |
| --- | --- | --- | --- | --- | --- | --- | --- | --- | --- | --- |
|  |  | HR (95% CI) | P-val | Q-val | HR (95% CI) | P-val | Q-val | HRR (CI) | P-val | Q-val |
| O02 | Other abnormal products of conception | 1.06 (1.01-1.13) | 0.027 | 0.069 | 0.96 (0.90-1.02) | 0.146 | 0.271 | 1.11 (1.03-1.21) | 9.83E-03 | 0.00 |
| O14 | Pre-eclampsia | 1.00 (0.93-1.08) | 0.961 | 0.961 | 1.07 (0.99-1.16) | 0.088 | 0.191 | 0.93 (0.83-1.04) | 0.231 | 0.43 |
| O24 | Diabetes in pregnancy, childbirth, and the puerperium | 1.05 (0.95-1.14) | 0.337 | 0.406 | 1.06 (0.97-1.17) | 0.206 | 0.324 | 0.98 (0.86-1.12) | 0.783 | 0.92 |
| O28 | Abnormal findings on antenatal screening of mother | 0.89 (0.79-1.01) | 0.062 | 0.117 | 0.90 (0.79-1.02) | 0.107 | 0.218 | 0.99 (0.83-1.19) | 0.911 | 0.97 |
| O30 | Multiple gestation | 0.89 (0.82-0.97) | 6.47E-03 | 0.0190 | 1.04 (0.96-1.12) | 0.348 | 0.460 | 0.86 (0.77-0.96) | 8.57E-03 | 0.00 |
| O35 | Maternal care for known or suspected fetal abnormality and damage | 0.89 (0.77-1.04) | 0.134 | 0.213 | 1.06 (0.92-1.22) | 0.432 | 0.536 | 0.84 (0.69-1.03) | 0.092 | 0.25 |
| O36 | Maternal care for other fetal problems | 0.95 (0.90-1.01) | 0.105 | 0.180 | 0.94 (0.89-1.00) | 0.061 | 0.164 | 1.01 (0.93-1.10) | 0.820 | 0.93 |
| O42 | Premature rupture of membranes | 1.04 (0.98-1.10) | 0.202 | 0.286 | 0.94 (0.88-1.00) | 0.052 | 0.151 | 1.11, (1.01-1.21) | 0.024 | 0.11 |
| O47 | False labor | 1.11 (1.04-1.18) | 1.27E-03 | 0.007 | 0.91 (0.85-0.98) | 8.05E-03 | 0.037 | 1.22 (1.11-1.34) | 4.31E-05 | 0.00 |
| O62 | Abnormalities of forces of labor | 0.94 (0.91-0.98) | 2.67E-03 | 0.012 | 0.94 (0.91-0.98) | 5.13E-03 | 0.026 | 1.00 (0.95-1.06) | 0.956 | 0.97 |
| O68 | Labor and delivery comp by abnormality of fetal acid-base balance | 0.97 (0.93-1.02) | 0.237 | 0.314 | 1.00 (0.95-1.04) | 0.904 | 0.904 | 0.98 (0.92-1.04) | 0.467 | 0.73 |
| O72 | Postpartum hemorrhage | 0.90 (0.86-0.93) | 1.94E-07 | 3.06E-06 | 0.89 (0.85-0.93) | 1.24E-07 | 2.54E-06 | 1.01 (0.95-1.07) | 0.786 | 0.92 |
| O82 | Encounter for cesarean delivery without indication | 1.03 (0.98-1.07) | 0.253 | 0.324 | 1.02 (0.98-1.07) | 0.307 | 0.434 | 1.00 (0.94-1.07) | 0.974 | 0.97 |
| O99 | Other maternal diseases classified elsewhere but complicating pregnancy / childbirth | 1.00 (0.96-1.04) | 0.910 | 0.933 | 1.02 (0.98-1.07) | 0.345 | 0.460 | 0.98 (0.93-1.04) | 0.525 | 0.74 |
| S02 | Fracture of skull and facial bones | 1.17 (0.99-1.39) | 0.063 | 0.117 | 0.84 (0.67-1.07) | 0.154 | 0.273 | 1.39 (1.04-1.86) | 0.026 | 0.11 |
| S05 | Injury of eye and orbit | 1.27 (1.14-1.42) | 1.25E-05 | 8.52E-05 | 1.11 (0.98-1.26) | 0.088 | 0.191 | 1.14 (0.97-1.35) | 0.115 | 0.25 |
| S93 | Disloc & sprain of joints & ligaments at ankle, foot & toe level | 1.12 (1.06-1.18) | 1.49E-04 | 8.74E-04 | 1.10 (1.04-1.17) | 1.29E-03 | 0.009 | 1.01 (0.93-1.10) | 0.768 | 0.92 |
| T14 | Injury of unspecified body region | 1.04 (0.93-1.17) | 0.453 | 0.516 | 1.14 (1.01-1.29) | 0.033 | 0.107 | 0.92 (0.78-1.08) | 0.305 | 0.52 |
| T43 | Poisoning by psychotropic drugs, not elsewhere classified | 1.32 (1.02-1.70) | 0.035 | 0.083 | 1.49 (1.15-1.95) | 2.91E-03 | 0.017 | 0.88 (0.61-1.27) | 0.495 | 0.74 |
| Z37 | Outcome of delivery | 0.74 (0.60-0.90) | 3.29E-03 | 0.012 | 0.96 (0.74-1.24) | 0.748 | 0.828 | 0.77 (0.55-1.07) | 0.123 | 0.25 |
| C50 | Malignant neoplasm of breast | 1.09 (0.90-1.32) | 0.401 | 0.470 | 0.85 (0.68-1.07) | 0.174 | 0.285 | 1.27 (0.94-1.72) | 0.117 | 0.25 |
| E66 | Overweight and obesity | 1.10 (1.06-1.15) | 6.00E-06 | 4.92E-05 | 1.04 (0.99-1.09) | 0.160 | 0.273 | 1.06 (1.00-1.13) | 0.060 | 0.22 |
| F10 | Alcohol related disorders | 1.11 (0.95-1.30) | 0.188 | 0.275 | 1.38 (1.16-1.63) | 2.46E-04 | 0.002 | 0.81 (0.64-1.02) | 0.075 | 0.23 |
| F20 | Schizophrenia | 0.94 (0.68-1.30) | 0.711 | 0.788 | 1.38 (0.98-1.94) | 0.065 | 0.164 | 0.68 (0.43-1.08) | 0.104 | 0.25 |
| F31 | Bipolar disorder | 1.30 (1.02-1.67) | 0.037 | 0.083 | 1.23 (0.94-1.59) | 0.125 | 0.245 | 1.06 (0.74-1.53) | 0.740 | 0.92 |
| F32 | Major depressive disorder, single episode | 1.14 (1.04-1.24) | 3.98E-03 | 0.014 | 1.06 (0.95-1.18) | 0.303 | 0.434 | 1.08 (0.94-1.23) | 0.285 | 0.50 |
| **F33** | **Major depressive disorder, recurrent** | **1.16 (1.05-1.27)** | **2.95E-03** | **0.012** | 0.97 (0.86-1.09) | 0.606 | 0.690 | 1.19 (1.02-1.39) | 0.025 | 0.17 |
| F34 | Persistent mood [affective] disorders | 0.97 (0.70-1.35) | 0.846 | 0.913 | 1.14 (0.79-1.66) | 0.474 | 0.556 | 0.85 (0.51-1.39) | 0.508 | 0.74 |
| **F41** | **Other anxiety disorders** | **1.31(1.18-1.45)** | **2.24E-07** | **3.06E-06** | 1.02 (0.89-1.16) | 0.825 | 0.867 | **1.29 (1.08-1.53)** | **3.87E-03** | **0.07** |
| F42 | Obsessive-compulsive disorder | 1.17(0.91-1.50) | 0.226 | 0.309 | 1.12 (0.83-1.52) | 0.453 | 0.546 | 1.04 (0.70-1.54) | 0.853 | 0.94 |
| F43 | Reaction to severe stress, and adjustment disorders | 1.08 (1.00-1.17) | 0.046 | 0.098 | 1.08 (0.99-1.18) | 0.068 | 0.164 | 1.00 (0.89-1.12) | 0.977 | 0.97 |
| **F50** | **Eating disorders** | **1.36 (1.10-1.69)** | **4.58E-03** | **0.014** | 0.98 (0.74-1.30) | 0.895 | 0.904 | 1.39 (0.97-1.98) | 0.071 | 0.23 |
| **F60** | **Specific personality disorders** | **1.34 (1.20-1.50)** | **4.50E-07** | **4.61E-06** | **1.40 (1.23-1.58)** | **2.70E-07** | **3.68E-06** | 0.96 (0.81-1.14) | 0.673 | 0.89 |
| **F99** | **Mental disorder, not otherwise specified** | 1.15, (0.94-1.40) | 0.181 | 0.275 | **1.29 (1.04-1.60)** | **0.019** | **0.064** | 0.89 (0.66-1.20) | 0.438 | 0.77 |
| **G40** | **Epilepsy and recurrent seizures** | **1.24 (1.05-1.46)** | **0.011** | **0.031** | **1.59 (1.35-1.88)** | **4.29E-08** | **1.76E-06** | **0.78 (0.62-0.98)** | **0.032** | **0.13** |
| I10 | Essential (primary) hypertension | 1.11 (1.00-1.24) | 0.057 | 0.117 | **1.16 (1.03-1.29)** | **0.011** | **0.044** | 0.96 (0.82-1.12) | 0.602 | 0.84 |
| I26 | Pulmonary embolism | 1.21 (0.95-1.54) | 0.124 | 0.203 | 0.96 (0.72-1.27) | 0.777 | 0.839 | 1.26 (0.86-1.84) | 0.232 | 0.43 |
| **J45** | **Asthma** | 1.05 (0.96-1.16) | 0.278 | 0.345 | **1.26 (1.14-1.39)** | **3.79E-06** | **3.88E-05** | **0.84 (0.73-0.96)** | **0.012** | **0.08** |
| **M22** | **Disorder of patella** | 1.01 (0.89-1.15) | 0.870 | 0.914 | **1.18 (1.03-1.34)** | **0.014** | **0.051** | 0.86 (0.71-1.03) | 0.107 | 0.23 |
| **M35** | **Other systemic involvement of connective tissue** | **1.58 (1.34-1.86)** | **5.08E-08** | **2.08E-06** | 1.13 (0.92-1.39) | 0.248 | 0.376 | **1.39 (1.08-1.80)** | **0.012** | **0.08** |
| N94 | Pain and other conditions assoc w/ female genital organs and menstrual cycle | 1.09 (0.99-1.20) | 0.090 | 0.160 | 0.95 (0.85-1.07) | 0.394 | 0.505 | 1.14 (0.99-1.33) | 0.081 | 0.23 |

**Supplemental Table 3.** Associations between index mother’s diagnosis and autism risk in maternal parallel cousins (HR, 95% CI), without (non-adjusted, main analysis) and after adjustment for the corresponding diagnosis in cousin’s mother (adjusted).

|  |  | Maternal parallel cousins – non-adjusted |  | Maternal parallel cousins –adjusted |  |
| --- | --- | --- | --- | --- | --- |
|  |  | HR (95% CI) | P-val | HR (95% CI) | P-val |
| O02 | Other abnormal products of conception | 1.06 (1.01-1.13) | 0.027 | 1.06 (1.01-1.13) | 0.028 |
| O14 | Pre-eclampsia | 1.00 (0.93-1.08) | 0.961 | 0.99 (0.91-1.07) | 0.738 |
| O24 | Diabetes in pregnancy, childbirth, and the puerperium | 1.05 (0.95-1.14) | 0.337 | 1.02 (0.93-1.12) | 0.638 |
| O28 | Abnormal findings on antenatal screening of mother | 0.89 (0.79-1.01) | 0.062 | 0.89 (0.79-1.01) | 0.066 |
| O30 | Multiple gestation | 0.89 (0.82-0.97) | 6.47E-03 | 0.90 (0.83-0.97) | 7.62E-03 |
| O35 | Maternal care for known or suspected fetal abnormality and damage | 0.89 (0.77-1.04) | 0.134 | 0.90 (0.77-1.04) | 0.158 |
| O36 | Maternal care for other fetal problems | 0.95 (0.90-1.01) | 0.105 | 0.95, (0.90-1.01) | 0.079 |
| O42 | Premature rupture of membranes | 1.04 (0.98-1.10) | 0.202 | 1.04 (0.98-1.10) | 0.214 |
| O47 | False labor | 1.11 (1.04-1.18) | 1.27E-03 | 1.10 (1.03-1.17) | 3.89E-03 |
| O62 | Abnormalities of forces of labor | 0.94 (0.91-0.98) | 2.67E-03 | 0.94 (0.91-0.98) | 1.25E-03 |
| O68 | Labor and delivery comp by abnormality of fetal acid-base balance | 0.97 (0.93-1.02) | 0.237 | 0.97 (0.93-1.01) | 0.179 |
| O72 | Postpartum hemorrhage | 0.90 (0.86-0.93) | 1.94E-07 | 0.90 (0.86-0.94) | 3.65E-07 |
| O82 | Encounter for cesarean delivery without indication | 1.03 (0.98-1.07) | 0.253 | 1.02 (0.98-1.07) | 0.350 |
| O99 | Other maternal diseases classified elsewhere but complicating pregnancy / childbrth | 1.00 (0.96-1.04) | 0.910 | 0.99 (0.95-1.03) | 0.550 |
| S02 | Fracture of skull and facial bones | 1.17 (0.99-1.39) | 0.063 | 1.16 (0.98-1.38) | 0.077 |
| S05 | Injury of eye and orbit | 1.27 (1.14-1.42) | 1.25E-05 | 1.26 (1.13-1.40) | 3.52E-05 |
| S93 | Disloc & sprain of joints & ligaments at ankl, ft & toe lev | 1.12 (1.06-1.18) | 1.49E-04 | 1.10 (1.04-1.17) | 7.68E-04 |
| T14 | Injury of unspecified body region | 1.04 (0.93-1.17) | 0.453 | 1.03 (0.92-1.16) | 0.571 |
| T43 | Psychotropic drugs, not elsewhere classified | 1.32 (1.02-1.70) | 0.035 | 1.30 (1.01-1.68) | 0.045 |
| Z37 | Outcome of delivery | 0.74 (0.60-0.90) | 3.29E-03 | 0.74 (0.60-0.90) | 3.14E-03 |
| C50 | Malignant neoplasm of breast | 1.09 (0.90-1.32) | 0.401 | 1.08 (0.89-1.31) | 0.437 |
| E66 | Overweight and obesity | 1.10 (1.06-1.15) | 6.00E-06 | 1.08 (1.03-1.12) | 7.27E-04 |
| F10 | Alcohol related disorders | 1.11 (0.95-1.30) | 0.188 | 1.09 (0.93-1.28) | 0.275 |
| F20 | Schizophrenia | 0.94 (0.68-1.30) | 0.711 | 0.92 (0.67-1.28) | 0.636 |
| F31 | Bipolar disorder | 1.30 (1.02-1.67) | 0.037 | 1.29 (1.01-1.66) | 0.043 |
| F32 | Major depressive disorder, single episode | 1.14 (1.04-1.24) | 3.98E-03 | 1.12 (1.02-1.22) | 0.013 |
| F33 | Major depressive disorder, recurrent | 1.16 (1.05-1.27) | 2.95E-03 | 1.13 (1.03-1.25) | 0.009 |
| F34 | Persistent mood [affective] disorders | 0.97 (0.70-1.35) | 0.846 | 0.95 (0.68-1.32) | 0.749 |
| F41 | Other anxiety disorders | 1.31(1.18-1.45) | 2.24E-07 | 1.28 (1.16-1.42) | 1.36E-06 |
| F42 | Obsessive-compulsive disorder | 1.17(0.91-1.50) | 0.226 | 1.15 (0.90-1.48) | 0.271 |
| F43 | Reaction to severe stress, and adjustment disorders | 1.08 (1.00-1.17) | 0.046 | 1.07 (0.99-1.15) | 0.105 |
| F50 | Eating disorders | 1.36 (1.10-1.69) | 4.58E-03 | 1.32 (1.07-1.64) | 0.010 |
| F60 | Specific personality disorders | 1.34 (1.20-1.50) | 4.50E-07 | 1.30 (1.16-1.46) | 5.96E-06 |
| F99 | Mental disorder, not otherwise specified | 1.15, (0.94-1.40) | 0.181 | 1.14 (0.93-1.39) | 0.205 |
| G40 | Epilepsy and recurrent seizures | 1.24 (1.05-1.46) | 0.011 | 1.23 (1.04-1.45) | 0.014 |
| I10 | Essential (primary) hypertension | 1.11 (1.00-1.24) | 0.057 | 1.09 (0.98-1.22) | 0.100 |
| I26 | Pulmonary embolism | 1.21 (0.95-1.54) | 0.124 | 1.20 (0.95-1.53) | 0.131 |
| J45 | Asthma | 1.05 (0.96-1.16) | 0.278 | 1.04 (0.94-1.14) | 0.455 |
| M22 | Disorder of patella | 1.01 (0.89-1.15) | 0.870 | 1.00 (0.88-1.15) | 0.963 |
| M35 | Other systemic involvement of connective tissue | 1.58 (1.34-1.86) | 5.08E-08 | 1.55 (1.32-1.83) | 1.23E-07 |
| N94 | Pain and other conditions assoc w/ female genital organs and menstrual cycle | 1.09 (0.99-1.20) | 0.090 | 1.08 (0.98-1.19) | 0.133 |

**Supplemental Table 4.** Fecundity (mean number of children) in index mother’s sisters and brothers, conditional on the presence of the given diagnosis in the index mother.

|  |  | Index mother's sister |  |  | Index mother's brother |  |  |
| --- | --- | --- | --- | --- | --- | --- | --- |
|  |  | # children (+) | # children (-) | Ratio (+)/(-) | # children (+) | # children (-) | Ratio (+)/(-) |
| O02 | Other abnormal products of conception | 1.76 | 1.69 | 1.04 | 1.47 | 1.42 | 1.04 |
| O14 | Pre-eclampsia | 1.64 | 1.70 | 0.96 | 1.36 | 1.43 | 0.95 |
| O24 | Diabetes in pregnancy, childbirth, and the puerperium | 1.70 | 1.70 | 1.00 | 1.42 | 1.42 | 0.99 |
| O28 | Abnormal findings on antenatal screening of mother | 1.66 | 1.70 | 0.97 | 1.33 | 1.43 | 0.93 |
| O30 | Multiple gestation | 1.76 | 1.70 | 1.04 | 1.48 | 1.42 | 1.04 |
| O35 | Maternal care for known or suspected fetal abnormality and damage | 1.62 | 1.70 | 0.95 | 1.33 | 1.43 | 0.93 |
| O36 | Maternal care for other fetal problems | 1.67 | 1.70 | 0.98 | 1.37 | 1.43 | 0.96 |
| O42 | Premature rupture of membranes | 1.66 | 1.71 | 0.97 | 1.36 | 1.43 | 0.95 |
| O47 | False labor | 1.67 | 1.70 | 0.98 | 1.35 | 1.43 | 0.94 |
| O62 | Abnormalities of forces of labor | 1.70 | 1.70 | 1.00 | 1.43 | 1.42 | 1.00 |
| O68 | Labor and delivery comp by abnormality of fetal acid-base balance | 1.64 | 1.72 | 0.95 | 1.37 | 1.44 | 0.95 |
| O72 | Postpartum hemorrhage | 1.46 | 1.84 | 0.79 | 1.16 | 1.58 | 0.74 |
| O82 | Encounter for cesarean delivery without indication | 1.71 | 1.70 | 1.00 | 1.44 | 1.42 | 1.01 |
| O99 | Other maternal diseases complicating pregnancy / childbirth | 1.61 | 1.74 | 0.92 | 1.31 | 1.47 | 0.89 |
| S02 | Fracture of skull and facial bones | 1.78 | 1.70 | 1.05 | 1.41 | 1.42 | 0.99 |
| S05 | Injury of eye and orbit | 1.74 | 1.70 | 1.03 | 1.46 | 1.42 | 1.03 |
| S93 | Disloc & sprain of joints & ligaments at ankl, ft & toe lev | 1.74 | 1.70 | 1.03 | 1.45 | 1.42 | 1.02 |
| T14 | Injury of unspecified body region | 1.73 | 1.70 | 1.02 | 1.44 | 1.42 | 1.01 |
| T43 | Psychotropic drugs, not elsewhere classified | 1.76 | 1.70 | 1.04 | 1.32 | 1.42 | 0.93 |
| Z37 | Outcome of delivery | 1.70 | 1.81 | 0.94 | 1.42 | 1.50 | 0.95 |
| C50 | Malignant neoplasm of breast | 1.86 | 1.70 | 1.10 | 1.63 | 1.42 | 1.15 |
| E66 | Overweight and obesity | 1.61 | 1.72 | 0.93 | 1.31 | 1.46 | 0.90 |
| F10 | Alcohol related disorders | 1.75 | 1.70 | 1.03 | 1.24 | 1.43 | 0.87 |
| F20 | Schizophrenia | 1.80 | 1.70 | 1.06 | 1.20 | 1.43 | 0.84 |
| F31 | Bipolar disorder | 1.71 | 1.70 | 1.01 | 1.33 | 1.42 | 0.93 |
| F32 | Major depressive disorder, single episode | 1.70 | 1.70 | 1.00 | 1.40 | 1.43 | 0.98 |
| F33 | Major depressive disorder, recurrent | 1.68 | 1.70 | 0.99 | 1.35 | 1.43 | 0.94 |
| F34 | Persistent mood [affective] disorders | 1.80 | 1.70 | 1.06 | 1.34 | 1.42 | 0.94 |
| F41 | Other anxiety disorders | 1.66 | 1.70 | 0.98 | 1.32 | 1.43 | 0.93 |
| F42 | Obsessive-compulsive disorder | 1.61 | 1.70 | 0.95 | 1.24 | 1.43 | 0.87 |
| F43 | Reaction to severe stress, and adjustment disorders | 1.72 | 1.70 | 1.01 | 1.38 | 1.43 | 0.97 |
| F50 | Eating disorders | 1.54 | 1.70 | 0.91 | 1.19 | 1.43 | 0.84 |
| F60 | Specific personality disorders | 1.65 | 1.70 | 0.97 | 1.23 | 1.43 | 0.86 |
| F99 | Mental disorder, not otherwise specified | 1.66 | 1.70 | 0.97 | 1.29 | 1.43 | 0.91 |
| G40 | Epilepsy and recurrent seizures | 1.75 | 1.70 | 1.03 | 1.41 | 1.42 | 0.99 |
| I10 | Essential (primary) hypertension | 1.88 | 1.70 | 1.11 | 1.60 | 1.42 | 1.13 |
| I26 | Pulmonary embolism | 1.75 | 1.70 | 1.03 | 1.48 | 1.42 | 1.04 |
| J45 | Asthma | 1.71 | 1.70 | 1.00 | 1.39 | 1.43 | 0.98 |
| M22 | Disorder of patella | 1.77 | 1.70 | 1.04 | 1.46 | 1.42 | 1.02 |
| M35 | Other systemic involvement of connective tissue | 1.78 | 1.70 | 1.04 | 1.52 | 1.42 | 1.07 |
| N94 | Pain and oth cond assoc w fem gntl org and menstrual cycle | 1.79 | 1.70 | 1.06 | 1.45 | 1.42 | 1.02 |

**Supplemental Table 5.** Associations between the index mother’s diagnosis and autism risk in maternal parallel and cross cousins (HR, 95% CI), in the main analyses and after restricting the exposure period to 5 years for all children in the cohort. The associations marked with NA could not be estimated due to an insufficient number of exposed children in the restricted exposure period.

|  |  | Main analysis |  | Sensitivity analysis (exposure period) |  |
| --- | --- | --- | --- | --- | --- |
|  |  | Maternal parallel cousins | Maternal cross cousins | Maternal parallel cousins | Maternal cross cousins |
|  |  | HR (95% CI) | HR (95% CI) | HR (95% CI) | HR (95% CI) |
| O02 | Other abnormal products of conception | 1.06 (1.01-1.13) | 0.96 (0.90-1.02) | 1.09 (1.02-1.17) | 0.96 (0.89-1.03) |
| O14 | Pre-eclampsia | 1.00 (0.93-1.08) | 1.07 (0.99-1.16) | 1.00 (0.92-1.09) | 1.11 (1.01-1.21) |
| O24 | Diabetes in pregnancy, childbirth, and the puerperium | 1.05 (0.95-1.14) | 1.06 (0.97-1.17) | 1.06 (0.95-1.18) | 1.04 (0.92-1.17) |
| O28 | Abnormal findings on antenatal screening of mother | 0.89 (0.79-1.01) | 0.90 (0.79-1.02) | 0.95 (0.82-1.09) | 0.97 (0.84-1.14) |
| O30 | Multiple gestation | 0.89 (0.82-0.97) | 1.04 (0.96-1.12) | 0.90 (0.82-0.98) | 1.06 (0.97-1.16) |
| O35 | Maternal care for known or suspected fetal abnormality and damage | 0.89 (0.77-1.04) | 1.06 (0.92-1.22) | 0.88 (0.73-1.06) | 1.01 (0.84-1.21) |
| O36 | Maternal care for other fetal problems | 0.95 (0.90-1.01) | 0.94 (0.89-1.00) | 0.97 (0.91-1.03) | 0.95 (0.88-1.02) |
| O42 | Premature rupture of membranes | 1.04 (0.98-1.10) | 0.94 (0.88-1.00) | 1.03 (0.96-1.09) | 0.95, (0.89-1.03) |
| O47 | False labor | 1.11 (1.04-1.18) | 0.91 (0.85-0.98) | 1.11 (1.04-1.20) | 0.94 (0.87-1.03) |
| O62 | Abnormalities of forces of labor | 0.94 (0.91-0.98) | 0.94 (0.91-0.98) | 0.96 (0.93-1.00) | 0.94 (0.90-0.98) |
| O68 | Labor and delivery comp by abnormality of fetal acid-base balance | 0.97 (0.93-1.02) | 1.00 (0.95-1.04) | 0.99 (0.94-1.03) | 1.00 (0.95-1.05) |
| O72 | Postpartum hemorrhage | 0.90 (0.86-0.93) | 0.89 (0.85-0.93) | 0.96 (0.91-1.00) | 0.93 (0.88-0.98) |
| O82 | Encounter for cesarean delivery without indication | 1.03 (0.98-1.07) | 1.02 (0.98-1.07) | 1.03 (0.99-1.08) | 1.04 (0.99-1.09) |
| O99 | Other maternal diseases classified elsewhere but complicating pregnancy / childbirth | 1.00 (0.96-1.04) | 1.02 (0.98-1.07) | 1.04 (1.00-1.09) | 1.05 (1.00-1.10) |
| S02 | Fracture of skull and facial bones | 1.17 (0.99-1.39) | 0.84 (0.67-1.07) | 1.29 (0.99-1.68) | 1.04 (0.74-1.47) |
| S05 | Injury of eye and orbit | 1.27 (1.14-1.42) | 1.11 (0.98-1.26) | 1.05 (0.86-1.29) | 1.17 (0.96-1.43) |
| S93 | Disloc & sprain of joints & ligaments at ankle, foot & toe level | 1.12 (1.06-1.18) | 1.10 (1.04-1.17) | 1.06 (0.97-1.15) | 1.11 (1.02-1.22) |
| T14 | Injury of unspecified body region | 1.04 (0.93-1.17) | 1.14 (1.01-1.29) | 1.15 (0.96-1.37) | 1.02 (0.83-1.25) |
| T43 | Poisoning by psychotropic drugs, not elsewhere classified | 1.32 (1.02-1.70) | 1.49 (1.15-1.95) | NA | NA |
| Z37 | Outcome of delivery | 0.74 (0.60-0.90) | 0.96 (0.74-1.24) | 0.80 (0.66-0.96) | 0.87 (0.71-1.08) |
| C50 | Malignant neoplasm of breast | 1.09 (0.90-1.32) | 0.85 (0.68-1.07) | NA | NA |
| E66 | Overweight and obesity | 1.10 (1.06-1.15) | 1.04 (0.99-1.09) | 1.07 (1.01-1.13) | 1.07 (1.00-1.13) |
| F10 | Alcohol related disorders | 1.11 (0.95-1.30) | 1.38 (1.16-1.63) | 1.08 (0.84-1.39) | 1.13 (0.84-1.50) |
| F20 | Schizophrenia | 0.94 (0.68-1.30) | 1.38 (0.98-1.94) | NA | NA |
| F31 | Bipolar disorder | 1.30 (1.02-1.67) | 1.23 (0.94-1.59) | NA | NA |
| F32 | Major depressive disorder, single episode | 1.14 (1.04-1.24) | 1.06 (0.95-1.18) | 1.14 (0.98-1.32) | 0.98 (0.81-1.18) |
| F33 | Major depressive disorder, recurrent | 1.16 (1.05-1.27) | 0.97 (0.86-1.09) | 1.12 (0.95-1.34) | 0.89 (0.71-1.12) |
| F34 | Persistent mood [affective] disorders | 0.97 (0.70-1.35) | 1.14 (0.79-1.66) | NA | NA |
| F41 | Other anxiety disorders | 1.31 (1.18-1.45) | 1.02 (0.89-1.16) | 0.97 (0.79-1.20) | 0.82 (0.62-1.07) |
| F42 | Obsessive-compulsive disorder | 1.17 (0.91-1.50) | 1.12 (0.83-1.52) | NA | NA |
| F43 | Reaction to severe stress, and adjustment disorders | 1.08 (1.00-1.17) | 1.08 (0.99-1.18) | 1.04 (0.91-1.19) | 1.15 (0.99-1.34) |
| F50 | Eating disorders | 1.36 (1.10-1.69) | 0.98 (0.74-1.30) | 1.32 (0.99-1.78) | 1.13 (0.78-1.64) |
| F60 | Specific personality disorders | 1.34 (1.20-1.50) | 1.40 (1.23-1.58) | 1.47 (1.24-1.74) | 1.23 (0.99-1.53) |
| F99 | Mental disorder, not otherwise specified | 1.15 (0.94-1.40) | 1.29 (1.04-1.60) | 0.76 (0.50-1.16) | 1.22 (0.81-1.84) |
| G40 | Epilepsy and recurrent seizures | 1.24 (1.05-1.46) | 1.59 (1.35-1.88) | 1.19 (0.97-1.48) | 1.31 (1.04-1.66) |
| I10 | Essential (primary) hypertension | 1.11 (1.00-1.24) | 1.16 (1.03-1.29) | 1.06 (0.85-1.33) | 1.22 (0.97-1.54) |
| I26 | Pulmonary embolism | 1.21 (0.95-1.54) | 0.96 (0.72-1.27) | NA | NA |
| J45 | Asthma | 1.05 (0.96-1.16) | 1.26 (1.14-1.39) | 0.95 (0.81-1.11) | 1.18 (1.01-1.38) |
| M22 | Disorder of patella | 1.01 (0.89-1.15) | 1.18 (1.03-1.34) | 0.96 (0.78-1.17) | 1.19 (0.97-1.44) |
| M35 | Other systemic involvement of connective tissue | 1.58 (1.34-1.86) | 1.13 (0.92-1.39) | 1.33 (0.97-1.81) | 0.87 (0.57-1.31) |
| N94 | Pain and other conditions assoc w/ female genital organs and menstrual cycle | 1.09 (0.99-1.20) | 0.95 (0.85-1.07) | 1.11 (0.95-1.30) | 1.09 (0.90-1.31) |

**Supplemental Table 6.** Associations between the index mother’s diagnosis and autism risk in maternal parallel and cross cousins (HR, 95% CI), in the main analyses and after removing families where the index mother has a diagnosis of autism.

|  |  | Main analysis |  | Sensitivity analysis (autism dx exclusion) |  |
| --- | --- | --- | --- | --- | --- |
|  |  | Maternal parallel cousins | Maternal cross cousins | Maternal parallel cousins | Maternal cross cousins |
|  |  | HR (95% CI) | HR (95% CI) | HR (95% CI) | HR (95% CI) |
| O02 | Other abnormal products of conception | 1.06 (1.01-1.13) | 0.96 (0.90-1.02) | 1.06 (1.01-1.13) | 0.95 (0.90-1.01) |
| O14 | Pre-eclampsia | 1.00 (0.93-1.08) | 1.07 (0.99-1.16) | 0.99 (0.91-1.07) | 1.05 (0.96-1.14) |
| O24 | Diabetes in pregnancy, childbirth, and the puerperium | 1.05 (0.95-1.14) | 1.06 (0.97-1.17) | 1.03 (0.94-1.13) | 1.06 (0.96-1.17) |
| O28 | Abnormal findings on antenatal screening of mother | 0.89 (0.79-1.01) | 0.90 (0.79-1.02) | 0.90 (0.80-1.02) | 0.86 (0.75-0.98) |
| O30 | Multiple gestation | 0.89 (0.82-0.97) | 1.04 (0.96-1.12) | 0.90 (0.83-0.97) | 1.02 (0.94-1.11) |
| O35 | Maternal care for known or suspected fetal abnormality and damage | 0.89 (0.77-1.04) | 1.06 (0.92-1.22) | 0.89 (0.76-1.04) | 1.04 (0.90-1.20) |
| O36 | Maternal care for other fetal problems | 0.95 (0.90-1.01) | 0.94 (0.89-1.00) | 0.96 (0.90-1.02) | 0.95 (0.89-1.01) |
| O42 | Premature rupture of membranes | 1.04 (0.98-1.10) | 0.94 (0.88-1.00) | 1.04 (0.98-1.10) | 0.94 (0.88-1.00) |
| O47 | False labor | 1.11 (1.04-1.18) | 0.91 (0.85-0.98) | 1.10 (1.03-1.17) | 0.91 (0.84-0.97) |
| O62 | Abnormalities of forces of labor | 0.94 (0.91-0.98) | 0.94 (0.91-0.98) | 0.95 (0.91-0.98) | 0.93 (0.89-0.97) |
| O68 | Labor and delivery comp by abnormality of fetal acid-base balance | 0.97 (0.93-1.02) | 1.00 (0.95-1.04) | 0.98 (0.94-1.02) | 1.00 (0.95-1.05) |
| O72 | Postpartum hemorrhage | 0.90 (0.86-0.93) | 0.89 (0.85-0.93) | 0.89 (0.86-0.93) | 0.88 (0.84-0.91) |
| O82 | Encounter for cesarean delivery without indication | 1.03 (0.98-1.07) | 1.02 (0.98-1.07) | 1.02 (0.97-1.06) | 1.01 (0.97-1.06) |
| O99 | Other maternal diseases classified elsewhere but complicating pregnancy / childbrth | 1.00 (0.96-1.04) | 1.02 (0.98-1.07) | 1.00 (0.96-1.04) | 1.02 (0.98-1.06) |
| S02 | Fracture of skull and facial bones | 1.17 (0.99-1.39) | 0.84 (0.67-1.07) | 1.19 (1.00-1.41) | 0.84 (0.66-1.06) |
| S05 | Injury of eye and orbit | 1.27 (1.14-1.42) | 1.11 (0.98-1.26) | 1.27 (1.14-1.41) | 1.13 (1.00-1.28) |
| S93 | Disloc & sprain of joints & ligaments at ankle, foot & toe level | 1.12 (1.06-1.18) | 1.10 (1.04-1.17) | 1.11 (1.05-1.18) | 1.11 (1.04-1.18) |
| T14 | Injury of unspecified body region | 1.04 (0.93-1.17) | 1.14 (1.01-1.29) | 1.05 (0.94-1.18) | 1.11 (0.98-1.25) |
| T43 | Poisoning by psychotropic drugs, not elsewhere classified | 1.32 (1.02-1.70) | 1.49 (1.15-1.95) | 1.29 (0.99-1.68) | 1.53 (1.17-1.99) |
| Z37 | Outcome of delivery | 0.74 (0.60-0.90) | 0.96 (0.74-1.24) | 0.80 (0.65-1.00) | 0.99 (0.76-1.29) |
| C50 | Malignant neoplasm of breast | 1.09 (0.90-1.32) | 0.85 (0.68-1.07) | 1.10 (0.91-1.33) | 0.85 (0.67-1.07) |
| E66 | Overweight and obesity | 1.10 (1.06-1.15) | 1.04 (0.99-1.09) | 1.10 (1.05-1.15) | 1.04 (0.99-1.09) |
| F10 | Alcohol related disorders | 1.11 (0.95-1.30) | 1.38 (1.16-1.63) | 1.12 (0.96-1.32) | 1.42 (1.20-1.69) |
| F20 | Schizophrenia | 0.94 (0.68-1.30) | 1.38 (0.98-1.94) | 0.94 (0.68-1.32) | 1.43 (1.02-2.01) |
| F31 | Bipolar disorder | 1.30 (1.02-1.67) | 1.23 (0.94-1.59) | 1.34 (1.04-1.71) | 1.23 (0.95-1.60) |
| F32 | Major depressive disorder, single episode | 1.14 (1.04-1.24) | 1.06 (0.95-1.18) | 1.11 (1.02-1.22) | 1.04 (0.93-1.16) |
| F33 | Major depressive disorder, recurrent | 1.16 (1.05-1.27) | 0.97 (0.86-1.09) | 1.15 (1.04-1.26) | 0.98 (0.87-1.11) |
| F34 | Persistent mood [affective] disorders | 0.97 (0.70-1.35) | 1.14 (0.79-1.66) | 0.96 (0.69-1.34) | 1.14 (0.78-1.66) |
| F41 | Other anxiety disorders | 1.31(1.18-1.45) | 1.02 (0.89-1.16) | 1.29 (1.16-1.43) | 1.02 (0.90-1.17) |
| F42 | Obsessive-compulsive disorder | 1.17(0.91-1.50) | 1.12 (0.83-1.52) | 1.20 (0.94-1.54) | 1.16 (0.85-1.57) |
| F43 | Reaction to severe stress, and adjustment disorders | 1.08 (1.00-1.17) | 1.08 (0.99-1.18) | 1.08 (0.99-1.16) | 1.07 (0.98-1.17) |
| F50 | Eating disorders | 1.36 (1.10-1.69) | 0.98 (0.74-1.30) | 1.31 (1.05-1.63) | 0.99 (0.74-1.32) |
| F60 | Specific personality disorders | 1.34 (1.20-1.50) | 1.40 (1.23-1.58) | 1.32 (1.18-1.48) | 1.40 (1.23-1.59) |
| F99 | Mental disorder, not otherwise specified | 1.15, (0.94-1.40) | 1.29 (1.04-1.60) | 1.16 (0.95-1.42) | 1.27 (1.02-1.58) |
| G40 | Epilepsy and recurrent seizures | 1.24 (1.05-1.46) | 1.59 (1.35-1.88) | 1.21 (1.02-1.43) | 1.58 (1.33-1.87) |
| I10 | Essential (primary) hypertension | 1.11 (1.00-1.24) | 1.16 (1.03-1.29) | 1.10 (0.98-1.22) | 1.17 (1.04-1.31) |
| I26 | Pulmonary embolism | 1.21 (0.95-1.54) | 0.96 (0.72-1.27) | 1.23 (1.00-1.57) | 0.94 (0.70-1.26) |
| J45 | Asthma | 1.05 (0.96-1.16) | 1.26 (1.14-1.39) | 1.05 (0.95-1.15) | 1.24 (1.12-1.37) |
| M22 | Disorder of patella | 1.01 (0.89-1.15) | 1.18 (1.03-1.34) | 1.01 (0.88-1.15) | 1.17 (1.02-1.33) |
| M35 | Other systemic involvement of connective tissue | 1.58 (1.34-1.86) | 1.13 (0.92-1.39) | 1.56 (1.32-1.84) | 1.16 (0.94-1.42) |
| N94 | Pain and other conditions assoc w/ female genital organs and menstrual cycle | 1.09 (0.99-1.20) | 0.95 (0.85-1.07) | 1.07 (0.97-1.18) | 0.97 (0.86-1.08) |

## Notes

### Competing Interest Statement

VK is currently employed by Takeda Pharmaceuticals, outside of submitted work

### Funding Statement

This work was supported by grants from the National Institute of Mental Health (MH124817, MH122394), and the Lundbeck Foundation (iPSYCH, Grant numbers R102-A9118 and R155-2014-1724).

